# PRISM: Phase-Resolved Isotropic Subtraction Mapping for Automated Multi-Phase CT Digital Subtraction Angiography

**DOI:** 10.64898/2026.09.10.26362169

**Authors:** Ronald Rodriguez, Ramiro Ramirez, Heena Rathore

**Affiliations:** Department of Urology and Medical Education UT Health San Antonio, San Antonio, TX 78229, US; Department of Computer Science, Texas State University San Marcos, TX 78666

**Keywords:** digital subtraction angiography, multi-phase CT, renal cell carcinoma, B-spline deformable registration, deep learning interpolation, parameter optimization

## Abstract

Multi-phase contrast-enhanced computed tomography (CT) is the gold standard for renal cell carcinoma (RCC) characterization, yet clinical interpretation relies on subjective visual comparison across phases. We present PRISM (Phase-Resolved Isotropic Subtraction Mapping), an open-source automated pipeline that transforms multi-phase CT acquisitions into registered digital subtraction angiography (DSA) volumes with color-coded enhancement maps. PRISM integrates six sequential processing stages: (1) DICOM loading with automated contrast-phase classification, (2) deep learning-based isotropic interpolation via RIFE, (3) automated kidney segmentation using TotalSegmentator v2, (4) enhancement-based tissue detection, (5) three-step deformable registration (rigid, affine, B-spline) using SimpleITK, and (6) dual-channel digital subtraction visualization. We present a systematic parameter optimization study comprising 200 registrations that runs across five patients and five experiments. Key findings: We identify an efficient registration configuration combining a 40 mm B-spline grid (within 6% of the 30 mm quality optimum at 36% lower computational cost), 5% metric sampling (equivalent quality to 25% at 3.2× speedup), and a single-level multi-resolution pyramid (avoiding the 5.4× overhead of a 4,2 pyramid with no quality benefit); we show that registration quality is effectively independent of interpolation target spacing from 0.5–3.0 mm, enabling a coarse-register/fine-apply strategy that computes the full transform at 3.0 mm (approximately 4 minutes per phase) and applies it to 0.5 mm volumes for high-resolution visualization. We also determine that a 40 HU subtraction noise threshold optimally balances signal-to-noise ratio (2.00) against sensitivity (14.2% enhancing volume retained), with higher thresholds (60–80 HU) favoring specificity and lower thresholds (20 HU) favoring sensitivity.

## 1. Introduction

Renal cell carcinoma (RCC) accounts for approximately 90% of kidney cancers and represents one of the most common malignancies worldwide (Filho *et al*. 2025), ranking among the top ten cancers in both men and women in the United States (Siegel *et al*. 2026). Multi-phase contrast-enhanced CT remains the standard for RCC detection, characterization, and staging, providing information about tumor vascularity through noncontrast, arterial, and venous phase acquisitions (Israel and Bosniak 2005). Despite advances in CT technology, radiologists must mentally register and subtract volumes acquired at different breath-hold positions, a cognitive burden that contributes to inter-observer variability and may cause subtle enhancement patterns to be overlooked, particularly in small or iso-attenuating lesions (Kim *et al*. 2002). This highlights an unmet need for automated volumetric phase-to-phase digital subtraction specifically tailored to solid organ multi-phase CT (e.g., renal protocol scans).

Digital subtraction angiography (DSA) principles have long been applied in vascular imaging to isolate contrast enhancement from background anatomy (Kotre and Marshall 2001). Applying DSA to volumetric CT data presents substantial technical challenges: spatial misalignment from patient motion and breathing, anisotropic voxel spacing requiring interpolation, and the computational demands of deformable registration across large three-dimensional volumes. Prior approaches to “4D CT angiography” employed rapid successive acquisitions to simulate angiographic sequences (Kato *et al*. 2004) (Te Kiefte *et al*. 2025) but have not addressed automated phase-to-phase digital subtraction for clinical renal imaging.

We present Phase-Resolved Isotropic Subtraction Mapping (PRISM), an open-source automated pipeline that addresses these challenges through a six-stage processing architecture. Our contributions are threefold: (1) an open-source pipeline for automated multi-phase renal CT-DSA; (2) a 200-run parameter optimization study that provides broadly applicable configuration insights for B-spline registration in high-resolution abdominal CT; and (3) a coarse-register/fine-apply strategy that preserves sub-millimeter visualization while reducing dual-phase registration time by approximately 60%.

We show that, with appropriate configuration, PRISM achieves clinically feasible runtimes while maintaining registration quality independent of voxel resolution. PRISM is released as an open-source, modular system with a documented web API and containerized deployment, facilitating reproducibility, reuse, and integration into existing research and clinical workflows. Although we focus on renal multi-phase CT, the parameter insights regarding B-spline grid spacing, metric sampling, multi-resolution pyramid configuration, and voxel resolution are directly applicable to other high-resolution abdominal and thoracic CT deformable registration pipelines.

## 2. Related Work

### 2.1 CT Digital Subtraction Angiography

CT-DSA extends conventional projection DSA to volumetric data by subtracting a pre-contrast volume from post-contrast acquisitions. Dual-energy CT subtraction was demonstrated for bone removal in head and neck angiography (Thomas *et al*. 2010) and subsequently applied in low-dose subtraction CT angiography using rapid kV-switching (Ma *et al*. 2018). These approaches assumed rigid alignment between acquisitions, limiting their applicability to body regions subject to respiratory motion. Deep learning methods have also been applied to CT image reconstruction more broadly (Koetzier *et al*. 2023). However, prior CT-DSA work has not addressed fully automated, phase-to-phase volumetric subtraction for renal imaging under realistic respiratory motion and heterogeneous clinical acquisition protocols.

### 2.2 Deep Learning Frame Interpolation

Huang *et al*. (Huang *et al*. 2022) introduced Real-Time Intermediate Flow Estimation (RIFE), a neural network for efficient video frame interpolation through iterative optical flow refinement using a five-block encoder (IFNet) operating at scales [16, 8, 4, 2, 1]. While originally designed for natural video, RIFE’s ability to synthesize intermediate frames from adjacent images makes it well-suited for through-plane super-resolution in medical imaging, where slice thickness (3–5 mm) substantially exceeds in-plane resolution (0.5–1.0 mm). Gambini *et al*. (Gambini *et al*. 2024) demonstrated RIFE applied to 3D tomography across length scales including CT, and Wu *et al*. (Wu *et al*. 2022) applied related slice imputation strategies for anisotropic medical image segmentation.

### 2.3 Automated Segmentation and Deformable Registration

Wasserthal *et al*. (Wasserthal *et al*. 2023) developed TotalSegmentator, which provided whole-body segmentation of 104 anatomical structures using an nnU-Net architecture (Isensee *et al*. 2021). The KiTS challenge established benchmarks for kidney, tumor, and cyst segmentation in corticomedullary-phase CT, first through KiTS19 (Heller *et al*. 2021) and subsequently through KiTS21 (Heller *et al*. 2023). Recent methods using these benchmarks have continued to advance state-of-the-art performance (Gan *et al*. 2026).

B-spline free-form deformation (Rueckert *et al*. 1999), models were used with non-rigid transformations using a grid of control points with smooth interpolation. Multi-resolution strategies apply registration at progressively finer scales, though as we demonstrate quantitatively in this paper, the computational cost at fine resolutions becomes prohibitive for high-resolution CT volumes, motivating our investigation of optimal pyramid configurations. SimpleITK (Lowekamp *et al*. 2013) provides the registration framework used throughout this work. Automated contrast-phase identification in multi-phase CT has been approached via DICOM metadata analysis and image-based classification methods (Reis *et al*. 2024); PRISM combines DICOM metadata, series description keywords, acquisition timing, and Hounsfield unit histogram features into a four-signal fusion scheme. Furthermore, in contrast to end-to-end learned registration or direct subtraction networks, PRISM combines classical intensity-based deformable registration with deep learning-based through-plane interpolation, a design that emphasizes interpretability, modularity, and compatibility with existing clinical CT protocols.

## 3. The PRISM Pipeline

PRISM is implemented as a modular Python web application (FastAPI, (Ramírez 2019); HTMX; PyTorch) comprising six processing stages executed sequentially by a PipelineController. The system is publicly available at github.com/rrodrig30/prism. A more detailed software architecture description is provided in Appendix A.

### Stage 1: Phase Classification

A four-signal weighted fusion approach (DICOM contrast tag: weight 0.25; series description keywords: 0.30; acquisition timing: 0.20; Hounsfield unit histogram: 0.25) classifies each series as noncontrast, arterial, or venous. These weights were chosen empirically based on retrospective inspection of typical renal CT protocols. Signals are normalized to [0,1] and combined via weighted sum with greedy best-match assignment.

### Stage 2: Isotropic Interpolation

RIFE (Huang *et al*. 2022) converts anisotropic CT volumes to isotropic spacing (default: 0.5 mm) through through-plane slice synthesis. Input slices are padded to multiples of 128 pixels and normalized from HU [−1000, 2000] to [0, 1] for inference.

### Stage 3: Kidney Segmentation

TotalSegmentator v2 (Wasserthal *et al*. 2023) localizes kidneys from the interpolated noncontrast volume, generating bilateral masks for region-of-interest definition.

### Stage 4: Enhancing Tissue Detection

Enhancement thresholding identifies voxels whose HU increase between noncontrast and contrast phases exceeds a configurable threshold, extending segmentation boundaries to include perilesional enhancing tissue.

### Stage 5: Multi-Phase Registration

A three-step cascade (rigid → affine → B-spline deformable) aligns each contrast phase to the noncontrast baseline using SimpleITK (Lowekamp *et al*. 2013)and Mattes Mutual Information. ITK threads are explicitly limited to 16 via SimpleITK.ProcessObject.SetGlobalDefaultNumberOfThreads(16) to prevent memory exhaustion on multi-core systems. Quantitative parameter analysis is presented in Section 4.

### Stage 6 Digital Subtraction and Visualization

Voxel-wise subtraction isolates enhancement; a configurable noise threshold suppresses misregistration artifacts. Arterial enhancement maps to red and venous to blue in a dual-channel overlay on grayscale anatomy.

## 4. Parameter Optimization Study

This section describes a systematic five-experiment study designed to identify optimal registration and subtraction parameter settings for PRISM. Each experiment isolates a single parameter while holding all others at fixed baseline values, enabling unambiguous attribution of performance differences to the parameter under investigation. The study encompasses 200 total registration runs across five clinical renal CT datasets and approximately 25 GPU-hours of computation.

### 4.1 Study Design and Patient Population

Five patients with pathologically confirmed RCC, each with three-phase clinical CT acquisitions (noncontrast, arterial corticomedullary, venous nephrographic), yielded 10 registration pairs (2 contrast phases × 5 patients) per parameter setting.

CT volumes were acquired on clinical scanners at 512×512 matrix size. Original Z-axis spacing ranged from 2.0 to 5.0 mm; in-plane pixel spacing was 0.68–0.90 mm. After RIFE interpolation to 1.0 mm isotropic spacing (except Experiment D, which varied this parameter), volume dimensions ranged from 300×512×512 to 705×512×512 voxels (78.6–184.7 million voxels per volume). All analyses used de-identified patient codes (Patient 1–5).

Patient 3 required special handling. The arterial phase DICOM series contained non-uniform slice spacing (maximum gap 23.6 mm versus 2–5 mm for all other patients), producing RIFE interpolation artifacts and precluding reliable arterial subtraction. The initially selected venous series had an incompatible field of view (X-axis origin +188.4 mm versus −192.0 mm for the noncontrast reference); a replacement venous series (origin −230.0 mm) was used. These issues did not affect Experiments A–E, which used all available registration pairs for quality metric computation, but produced consistently lower MI improvement values for Patient 3 across all five experiments.

### 4.2 Registration Protocol and Quality Metrics

Each registration used three sequential steps. Step 1 (rigid, 6 DOF): Euler3D transform, RegularStepGradientDescent optimizer (learning rate 1.0, minimum step 0.001, 200 iterations), multi-resolution pyramid [4,2,1]. Step 2 (affine, 12 DOF): initialized from rigid result, RegularStepGradientDescent (learning rate 0.5, 100 iterations), pyramid [2,1]. Step 3 (B-spline deformable): cubic B-spline transform, L-BFGS-B optimizer (gradient tolerance 1e-5, 100 function evaluations, 5 correction vectors). All steps used Mattes Mutual Information (MI) (50 bins, random voxel sampling). The primary quality metric was MI improvement = MI_after_bspline − MI_before (because Mattes MI is minimized by the optimizer, more negative values indicate improved alignment). Registration efficiency was defined as |MI improvement| / total time in minutes.

All experiments were executed on an NVIDIA H100 NVL GPU (93.1 GB VRAM) with 72 CPU cores. ITK thread count was limited to 16. Total benchmark wall time was approximately 25 hours.

### 4.3 Experiment A: B-Spline Grid Spacing

Objective: Determine the optimal B-spline control point grid spacing balancing registration quality against computational cost. Five grid spacings were evaluated: 20, 30, 40, 50, and 60 mm. The number of free B-spline parameters ranged from 2,510 (60 mm) to 38,603 (20 mm), a 15.4-fold range.

Registration time decreased monotonically with increasing grid spacing (Figure 1A), from 23.0 ± 10.1 minutes at 20 mm to 5.3 ± 2.2 minutes at 60 mm. Paradoxically, the finest grid spacing (20 mm) did not yield the best registration quality (Figure 1B). The 30 mm grid achieved the largest mean MI improvement (−0.240), outperforming the 20 mm grid (−0.190) despite having 2.9× fewer parameters. This reflects premature L-BFGS-B termination at 20 mm: with 38,603 parameters and only 100 function evaluations, the optimizer cannot adequately explore the high-dimensional parameter space. The 40 mm grid (−0.226) achieved quality within 6% of the 30 mm optimum at 36% lower cost. Registration efficiency (|MI improvement|/minute) increased monotonically from 0.008/min at 20 mm to 0.038/min at 60 mm (Figure 1C).

**Figure 1.**
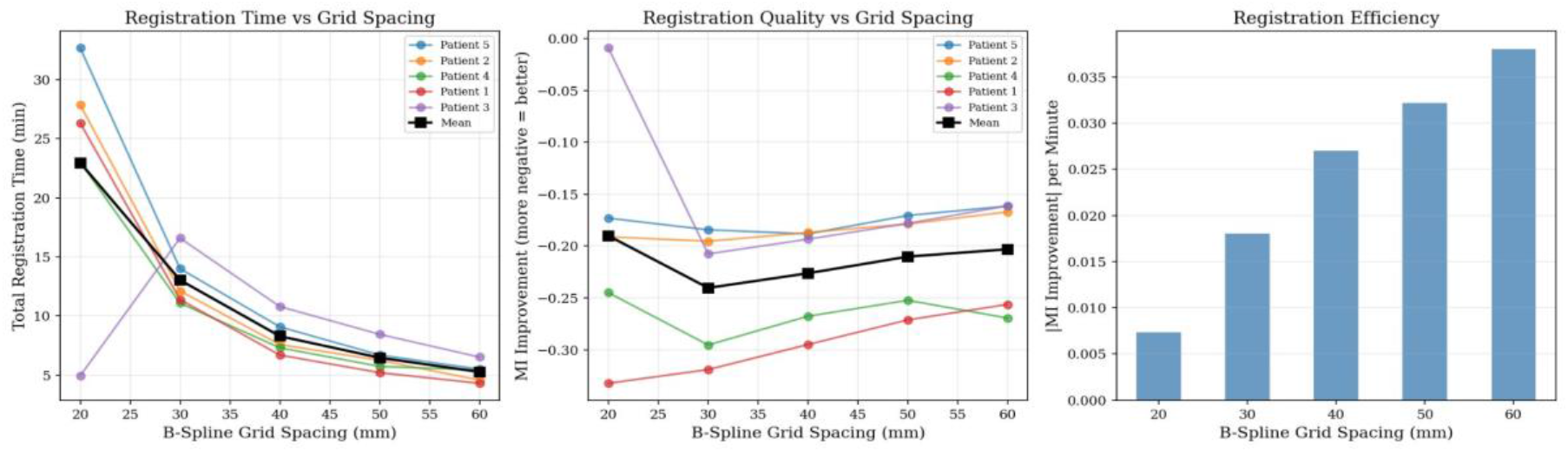
Effect of B-spline grid spacing on registration performance. (A) Total registration time (minutes) vs. grid spacing (20–60 mm) for each patient (colored lines) and cross-patient mean (bold black). (B) Registration quality vs. grid spacing. The 30 mm grid achieved the best mean MI improvement (−0.240); the 20 mm grid (−0.190) underperformed due to premature optimizer termination. (C) Registration efficiency (|MI improvement| per minute) by grid spacing, increasing monotonically from 0.008 to 0.038. Each data point: mean of 10 registration pairs.

**Table 1.** Registration performance by B-spline grid spacing. Mean ± SD across 10 registration pairs (2 phases × 5 patients) per setting. The 20 mm mean is attenuated by Patient 3, whose 60,306-parameter space caused immediate L-BFGS-B termination before any optimization could occur.

| Grid (mm) | B-Spline Params | Total Time (min) | B-Spline Time (min) | MI Improvement |
| --- | --- | --- | --- | --- |
| 20 | 38,603 | $23.0 \pm 10.1$ | $20.1 \pm 10.6$ | $-0.190 \pm 0.118$ |
| 30 | 13,222 | $13.0 \pm 7.0$ | $10.2 \pm 6.1$ | $-0.240 \pm 0.121$ |
| 40 | 6,545 | $8.3 \pm 4.3$ | $5.6 \pm 3.3$ | $-0.226 \pm 0.112$ |
| 50 | 4,054 | $6.5 \pm 3.2$ | $3.8 \pm 2.2$ | $-0.210 \pm 0.106$ |
| 60 | 2,510 | $5.3 \pm 2.2$ | $2.6 \pm 1.2$ | $-0.203 \pm 0.096$ |

Patient 3 exhibited a notable optimizer failure at 20 mm. With 60,306 parameters, nearly twice any other patient, L-BFGS-B terminated after 0 iterations because per-parameter gradient contributions were individually too small for the total gradient magnitude to exceed the fixed 1e-5 tolerance. At 30 mm (19,968 parameters), the same patient achieved the best arterial MI improvement in the cohort (−0.415), confirming that the registration was not unnecessary but that the convergence criterion was not scaled to the parameter space dimensionality. Taken together, these results illustrate that the optimizer budget must scale with the dimensionality of the B-spline parameter space; without sufficient evaluations, finer grids cannot realize their theoretical quality advantage.

A grid spacing of 40 mm was selected as the baseline for subsequent experiments, representing the best quality/speed tradeoff (MI improvement −0.226, total time 8.3 min, within 6% of the 30 mm quality optimum at 36% lower cost).

### 4.4 Experiment B: Metric Sampling Fraction

Objective: Determine the effect of metric voxel sampling density on registration quality and computation time. Four sampling fractions were evaluated: 5%, 10%, 15%, and 25%.

Registration time scaled approximately linearly with sampling fraction, from 4.1 ± 1.7 minutes at 5% to 13.2 ± 7.2 minutes at 25% (3.2× increase), consistent with the expected linear relationship between voxel count and metric evaluation cost (Figure 2A). In contrast, registration quality was remarkably insensitive to sampling density: MI improvement varied by less than 4% across the entire tested range (−0.221 at 5% versus −0.229 at 25%), with differences well within the inter-patient standard deviation of 0.08–0.12 (Figure 2B). This result indicates that 5% sampling captures sufficient spatial information for L-BFGS-B optimization at 40 mm grid spacing in abdominal CT. This demonstrates that relatively sparse metric sampling can be sufficient when the underlying deformation model is coarse, allowing substantial runtime reductions without sacrificing registration quality.

**Figure 2.**
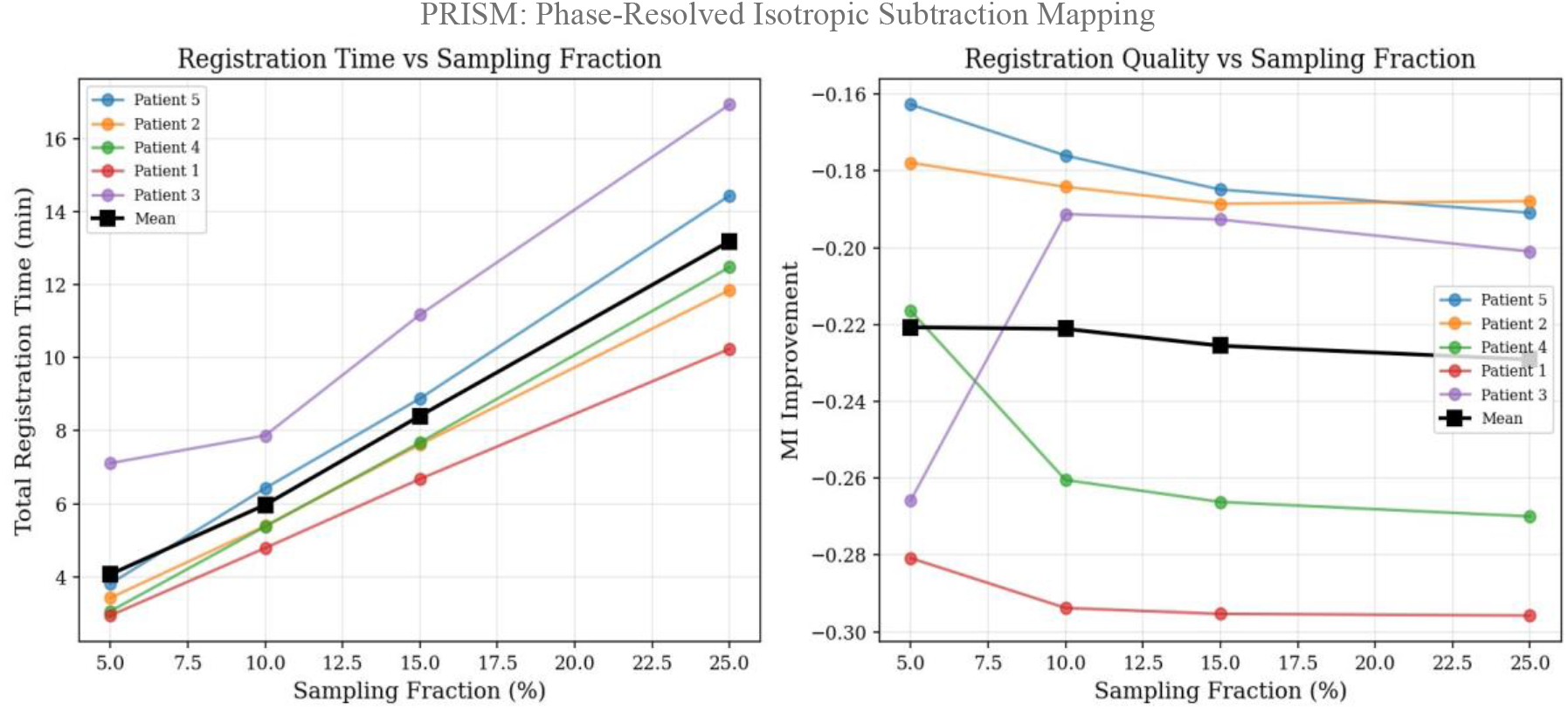
Effect of metric sampling fraction on registration performance. (A) Total registration time vs. sampling fraction (5–25%). Time scales approximately linearly (4.1 min at 5% to 13.2 min at 25%). (B) Registration quality (MI improvement) vs. sampling fraction. Quality varied by less than 4% across the full range (−0.221 at 5% vs. −0.229 at 25%), well within inter-patient variability. Each data point: mean of 10 registration pairs.

**Table 2.** Registration performance by metric sampling fraction. Mean ± SD across 10 registration pairs per setting. Grid spacing fixed at 40 mm.

| Sampling Fraction | Total Time (min) | B-Spline Time (min) | MI Improvement |
| --- | --- | --- | --- |
| 5% | 4.1 ± 1.7 | 2.5 | −0.221 ± 0.078 |
| 10% | 6.0 ± 2.9 | 3.9 | −0.221 ± 0.113 |
| 15% | 8.4 ± 4.5 | 5.6 | −0.226 ± 0.112 |
| 25% | 13.2 ± 7.2 | 9.1 | −0.229 ± 0.116 |

A sampling fraction of 5% is recommended for production use, achieving equivalent quality to the 15% baseline at 49% of computation time (4.1 versus 8.4 minutes).

### 4.5 Experiment C: Multi-Resolution Pyramid Configuration

Objective: Evaluate whether additional pyramid resolution levels improve B-spline registration quality. Three configurations were compared: single-level at 8× downsampling ([8], σ = 4 mm); single-level at 4× downsampling ([4], σ = 2 mm, baseline); two-level at 4× then 2× downsampling ([4,2], σ = 2 and 1 mm). The three configurations produced dramatically different computation times (Figure 3A). The [4,2] two-level pyramid required 45.6 ± 18.2 minutes—5.4× slower than [4] (8.4 ± 4.1 min) and 12.3× slower than [8] (3.7 ± 1.5 min). This extreme cost arose from the 2× refinement level, which ran B-spline optimization on 150–352 × 256 × 256 voxel images, requiring approximately 37 additional minutes per registration. Critically, the [4,2] configuration did not improve registration quality over the single-level [4] pyramid (−0.218 versus −0.225; Figure 3B). This counterintuitive result indicates that a 40 mm B-spline grid is too coarse to benefit from finer-resolution optimization: the control point spacing captures body-scale deformations already fully represented at 4× downsampling. The [8] configuration achieved lower quality (−0.158), a 30% relative reduction compared to [4], indicating that 8× downsampling discards too much spatial information for accurate metric evaluation. These findings indicate that multi-resolution pyramid design must be matched to the spatial scale of the deformation model; when the B-spline grid is coarse, additional fine-scale pyramid levels add cost without improving accuracy.

**Figure 3.**
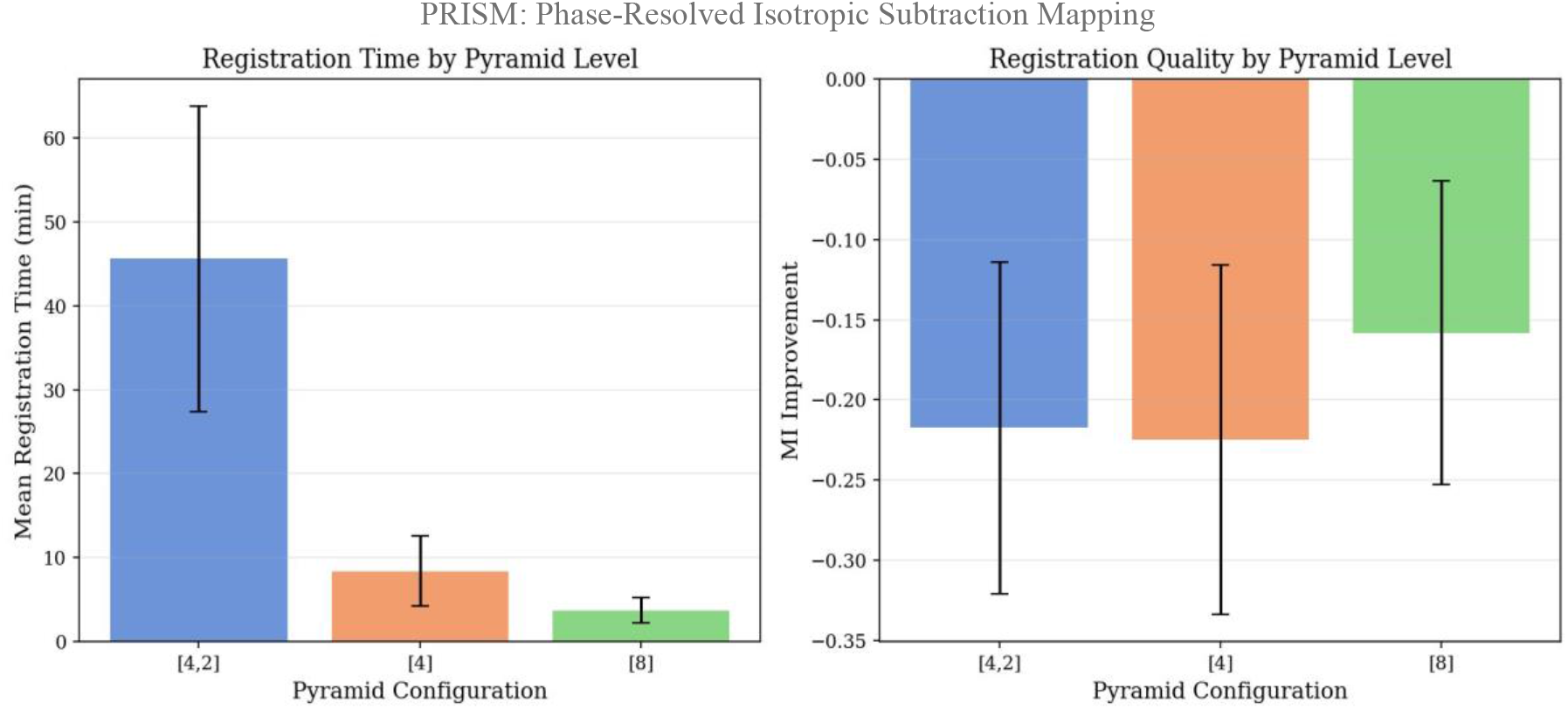
Effect of multi-resolution pyramid configuration on registration performance. (A) Mean registration time (±1 SD) for [4,2] (45.6 min), [4] (8.4 min), and [8] (3.7 min) configurations. (B) Mean MI improvement by pyramid configuration. The [4,2] pyramid (−0.218) did not improve quality over [4] (−0.225), confirming that 40 mm B-spline spacing is too coarse to benefit from finer-resolution optimization.

**Table 3.** Registration performance by multi-resolution pyramid configuration. Mean ± SD across 10 registration pairs per setting. Grid spacing 40 mm, sampling fraction 15%.

| Pyramid Config | Total Time (min) | B-Spline Time (min) | MI Improvement |
| --- | --- | --- | --- |
| [8] | 3.7 ± 1.5 | 1.0 | −0.158 ± 0.095 |
| [4] | 8.4 ± 4.1 | 5.6 | −0.225 ± 0.109 |
| [4,2] | 45.6 ± 18.2 | 42.8 | −0.218 ± 0.103 |

The single-level [4] pyramid is strongly recommended. The [4,2] configuration should be avoided, as it provides no quality benefit at 5.4× computational cost. The [8] configuration may suit rapid previews where 2.3× speedup is preferred over the 30% quality reduction.

### 4.6 Experiment D: Interpolation Target Spacing

Objective: Determine how pre-registration voxel resolution affects downstream registration quality and total pipeline time. For each target spacing (0.5, 1.0, 2.0, 3.0 mm isotropic), the full pipeline—RIFE interpolation, resampling to the noncontrast reference grid, and three-step registration—was re-executed independently.

**Table 4.**
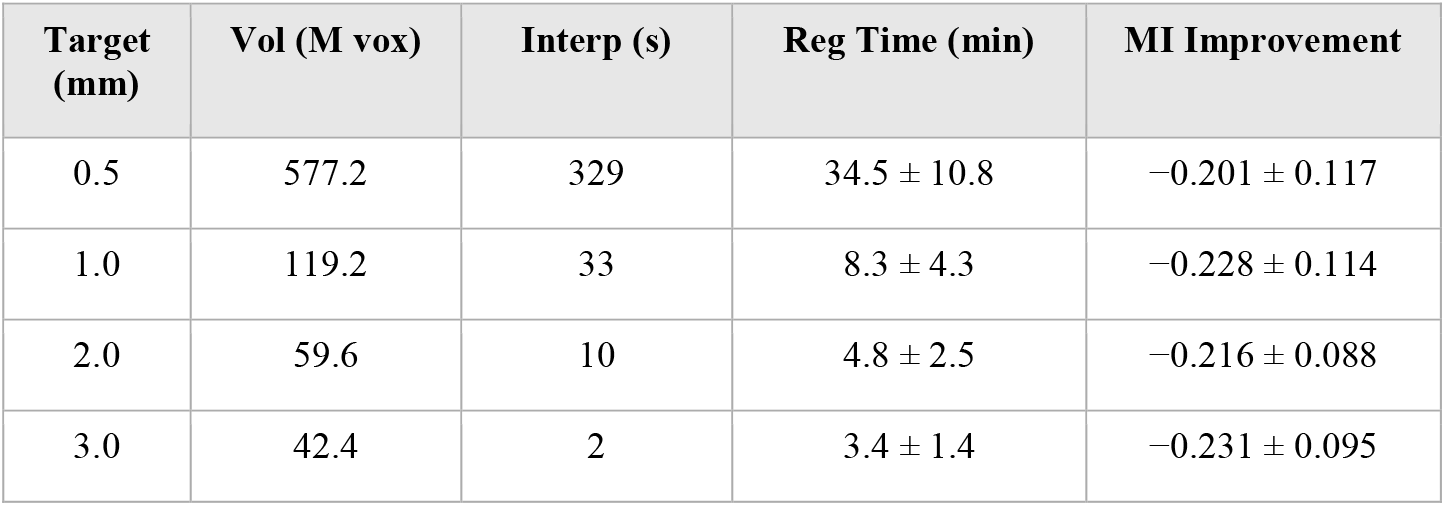
Pipeline performance by RIFE interpolation target spacing. Mean ± SD across 10 registration pairs per setting. Total pipeline time = RIFE interpolation + registration. The 0.5 mm runs completed after correction of a RIFE padding defect (32 vs. required 128 pixel multiples; see Appendix B).

Volume size scaled cubically with decreasing spacing: 0.5 mm volumes (577 M voxels) contained 4.8× more voxels than 1.0 mm (119 M) and 13.6× more than 3.0 mm (42 M; Figure 4C). Total pipeline time increased sharply at finer spacings: 40.0 minutes at 0.5 mm versus 8.8 minutes at 1.0 mm and 3.4 minutes at 3.0 mm (Figure 4A). Registration quality was essentially independent of interpolation target spacing across the full 0.5–3.0 mm range (Figure 4B): MI improvement ranged from −0.201 (0.5 mm) to −0.231 (3.0 mm), with differences not meaningful relative to the inter-patient standard deviation of 0.09–0.11. This confirms that B-spline registration at 40 mm grid spacing operates at a spatial scale far coarser than the voxel resolution; finer interpolation provides no additional information for the optimizer.

**Figure 4.**
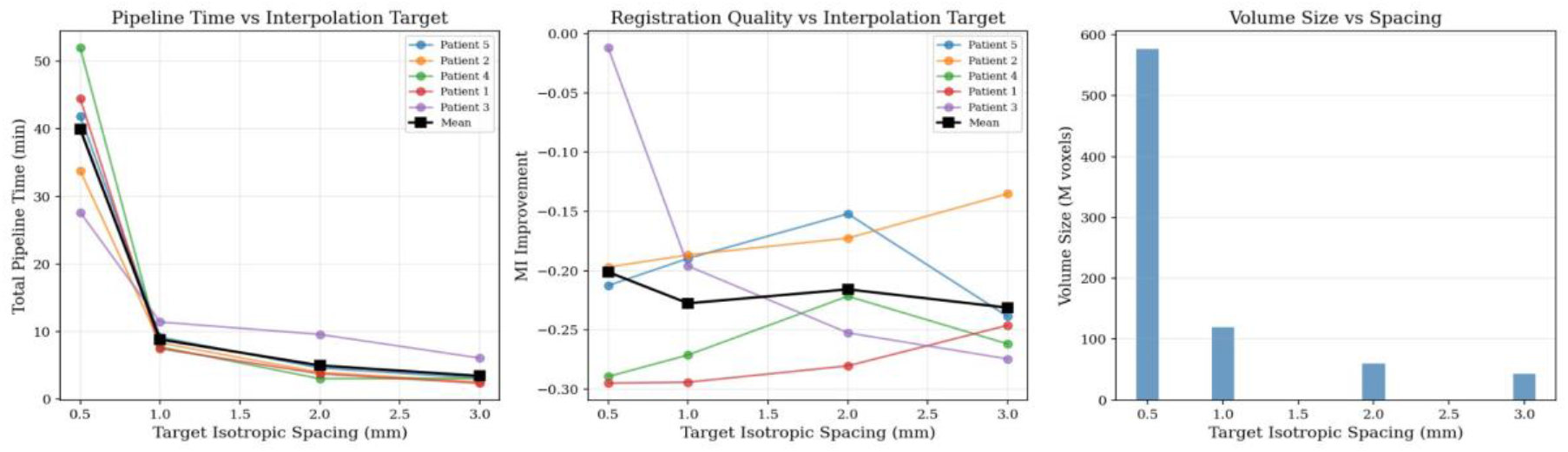
Effect of interpolation target spacing on pipeline performance. (A) Total pipeline time (RIFE + registration) vs. target isotropic spacing (0.5–3.0 mm): 40.0 min at 0.5 mm vs. 3.4 min at 3.0 mm. (B) Registration quality (MI improvement) is essentially constant across the full range. (C) Volume size (M voxels) by spacing: 0.5 mm volumes contain 4.8× more voxels than 1.0 mm.

These results directly motivate the **coarse-register/fine-apply strategy**: since registration quality is equivalent at 3.0 mm, the full rigid + affine + B-spline transform is computed at coarse resolution (~4 minutes/phase) and then applied to 0.5 mm volumes for downstream visualization and analysis. This decouples registration speed from output resolution, eliminating the 34.5-minute registration penalty at 0.5 mm while preserving sub-millimeter spatial detail for segmentation, subtraction, and 3D reconstruction

### 4.7 Experiment E: Subtraction Noise Threshold

Objective: Determine the optimal noise threshold for digital subtraction balancing signal-to-noise ratio (SNR) against sensitivity to true enhancing tissue. For each patient and contrast phase, a single registration was performed at default parameters (40 mm grid, 15% sampling, [4] pyramid), then subtraction was computed and each threshold (20, 40, 60, 80 HU) applied independently. SNR was computed as the mean enhancement in suprathreshold voxels divided by the standard deviation of the subtraction signal in non-enhancing body tissue.

**Table 5.** Subtraction quality by noise threshold. Mean ± SD across 10 registration pairs per setting.

| Threshold (HU) | SNR | Enhancing Vol (%) | Mean Enhancement (HU) |
| --- | --- | --- | --- |
| 20 | 1.53 ± 1.87 | 23.55 ± 10.2 | 199.9 |
| 40 | 2.00 ± 1.86 | 14.21 ± 9.3 | 233.1 |
| 60 | 2.48 ± 1.90 | 9.54 ± 9.4 | 269.0 |
| 80 | 3.02 ± 2.04 | 7.02 ± 9.3 | 310.8 |

SNR increased monotonically with noise threshold (Figure 5A), rising from 1.53 ± 1.87 at 20 HU to 3.02 ± 2.04 at 80 HU (97% improvement). Enhancing tissue volume decreased from 23.6% at 20 HU to 7.0% at 80 HU (Figure 5B), with the steepest reduction between 20 and 40 HU (9.3 percentage points), indicating that a large proportion of suprathreshold voxels at 20 HU represent misregistration noise. Mean enhancement in suprathreshold voxels increased from 200 HU to 311 HU across this range, confirming that higher thresholds selectively retain high-confidence enhancing voxels.

**Figure 5.**
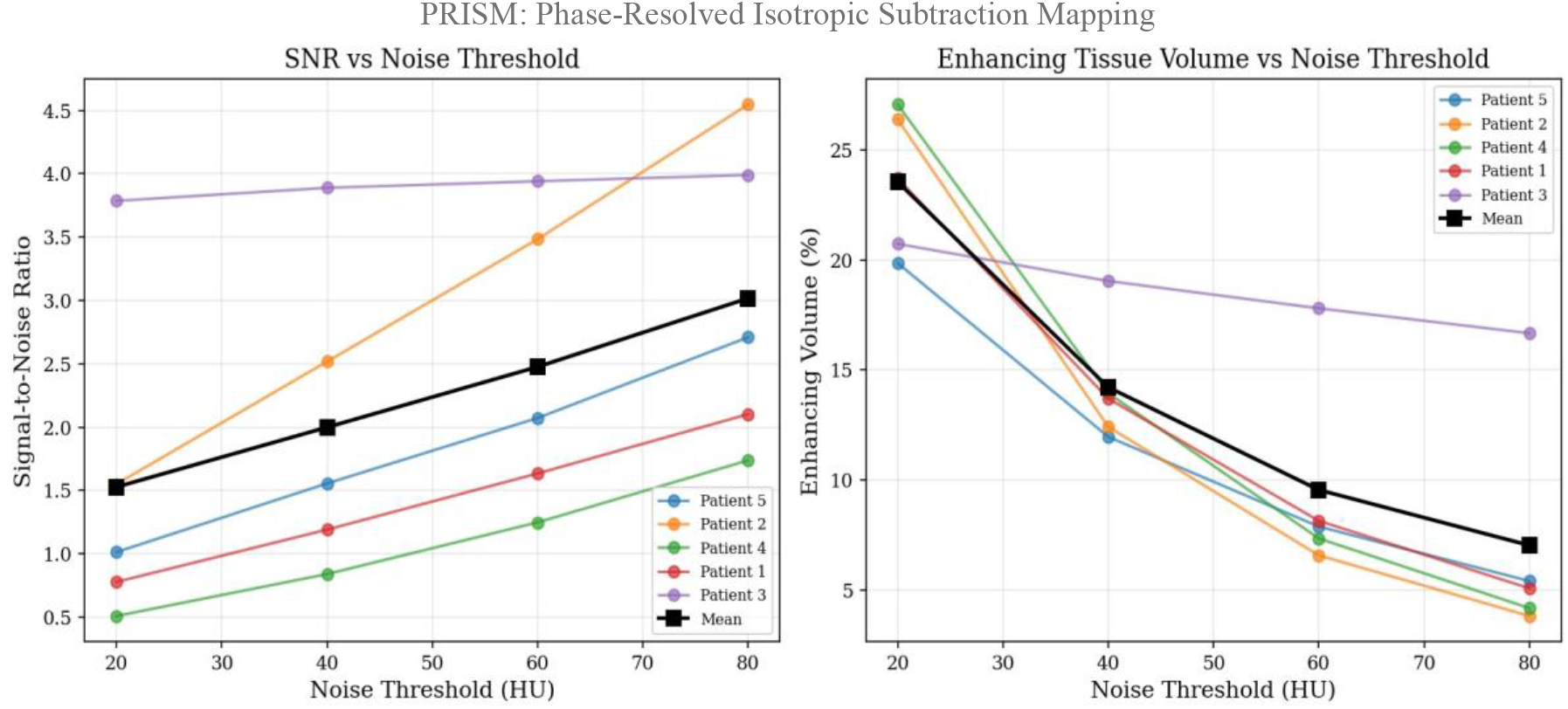
Effect of subtraction noise threshold on DSA quality. (A) SNR vs. noise threshold (20–80 HU): SNR increased from 1.53 to 3.02 (97% improvement). (B) Enhancing tissue volume (% of total voxels) vs. threshold; steepest reduction between 20 and 40 HU (9.3 percentage points). A threshold of 40 HU was selected as optimal, balancing SNR against sensitivity.

Inter-patient variability in SNR was substantial (SD 1.87–2.04), reflecting differences in contrast bolus timing, renal perfusion, and registration accuracy. A threshold of 40 HU is recommended, reducing apparent enhancing volume by 40% relative to 20 HU while retaining voxels with clinically meaningful enhancement (mean 233 HU, SNR 2.00). For high-specificity applications such as tumor delineation, a 60–80 HU threshold may be preferred; for high-sensitivity applications such as subtle enhancement detection, 20 HU is appropriate. Overall, these results emphasize that subtraction noise thresholds fundamentally trade sensitivity for visual SNR and therefore must be tuned to the specific clinical task and tolerance for false positives.

## 5. Optimized Configuration and End-to-End Results

### 5.1 Recommended Parameter Configuration

Table 6 summarizes the optimal parameters identified across all five experiments. The combined optimized configuration reduces dual-phase registration from the original ~20 minutes to approximately 8 minutes per study, a 60% reduction, with no meaningful quality compromise regardless of target output resolution.

**Table 6.** Summary of optimal parameters identified across all five experiments. *Registration quality is equivalent across 0.5–3.0 mm; the coarse-register/fine-apply strategy (Experiment D) enables 0.5 mm output with approximately 4 min/phase registration speed.

| Parameter | Tested Range | Optimal | Rationale | Relative time impact (H100 system) |
| --- | --- | --- | --- | --- |
| B-spline grid spacing | 20–60 mm | 30–40 mm | Best quality at 30 mm; best quality/speed at 40 mm for production | 40 mm yields ≈36% lower registration time than 30 mm. |
| Sampling fraction | 5–25% | 5% | Equivalent quality at 3.2× speedup vs. 15% baseline | 5% sampling gives ≈3.2× lower registration time than 25%. |
| Pyramid configuration | [8], [4], [4,2] | [4] | adds 5.4× cost with no quality improvement; [8] is faster but degrades quality. | [4,2] is ≈5.4× slower than the single-level [4] configuration, while [4] is ≈2.3× faster than [8] on our H100-based test system. |
| Interpolation target | 0.5–3.0 mm | 0.5 mm* | No registration quality difference; finest resolution for visualization | Total pipeline time grows sharply at finer spacing; 0.5 mm incurs ≈40 min vs. ≈3.4 min at 3.0 mm per study before coarse-register/fine-apply. |
| Noise threshold | 20–80 HU | 40 HU | Best sensitivity/specificity; 40% noise reduction vs. 20 HU | Negligible effect on registration time; affects visualization and post-processing only. |

\* Registration quality is equivalent across 0.5–3.0 mm. The coarse-register/fine-apply strategy computes the B-spline transform at 3.0 mm resolution (~4 minutes/phase) and applies the resolution-independent physical-coordinate transform to 0.5 mm volumes for downstream analysis.

With RIFE interpolation at 0.5 mm adding approximately 329 seconds per study, the total optimized pipeline time from DICOM load through registered 0.5 mm volumes is approximately 14 minutes on a single NVIDIA H100 GPU.

### 5.2 End-to-End Pipeline Demonstration

To demonstrate the complete pipeline with optimized parameters, five-panel composite kidney images were generated for all five patients. Automatic kidney slice selection used a bone valley detection algorithm: (1) the Gaussian-smoothed bone pixel count profile (σ = 3% of Z depth) was computed; (2) the local minimum in the central 25–75% of the Z-axis identified the lumbar/kidney region; (3) the slice with the best bilateral enhancement score within a ±5% window was selected. Body mask erosion (8 iterations for overlay, 5 for DSA panels) suppressed skin-edge misregistration artifacts. A physical Z-coordinate validity mask zeroed enhancement outside each contrast phase’s actual Z-axis coverage, handling field-of-view differences between acquisitions.

Figure 6 shows the five-panel composite for a representative patient (Patient 5). Panels display: (A) noncontrast grayscale anatomy; (B) arterial DSA (red on black); (C) venous DSA (blue on black); (D) dual-phase subtraction (arterial red + venous blue; overlap = purple/magenta); (E) dual-phase color overlay on noncontrast anatomy. Arterial enhancement was most prominent in the renal cortex and main renal arteries; venous enhancement showed diffuse parenchymal uptake. Effective color separation allowed immediate visual differentiation of vascular phases. Using the coarse-register/fine-apply strategy, total registration time for this patient was approximately 8 minutes for both contrast phases combined.

**Figure 6.**
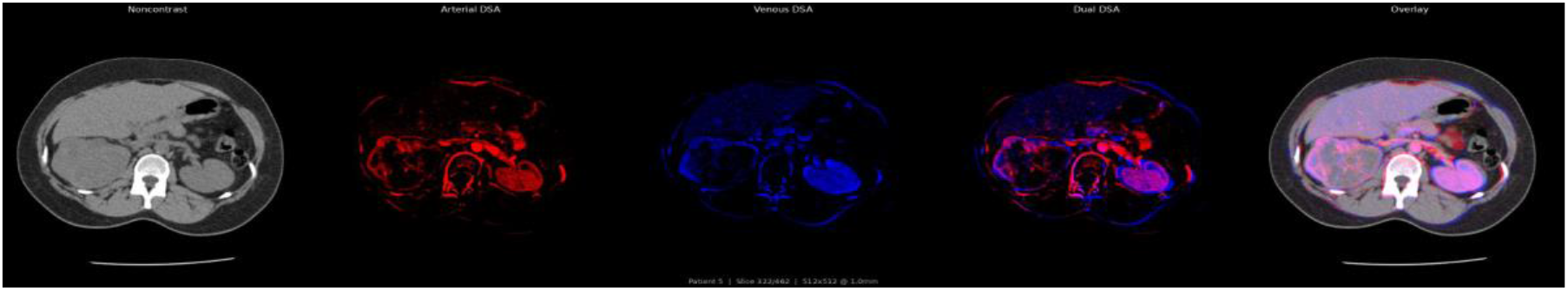
Five-panel composite output for Patient 5 at the automatically selected kidney axial level. (A) Noncontrast grayscale (window center 40 HU, width 400 HU). (B) Arterial DSA (red channel, black background). (C) Venous DSA (blue channel, black background). (D) Dual-phase subtraction (arterial red + venous blue; overlap = purple/magenta). (E) Dual-phase color overlay on noncontrast anatomy. Pipeline parameters: 0.5 mm interpolation, coarse-register at 3.0 mm/fine-apply at 1.0 mm, 40 mm B-spline grid, 5% sampling, [4] pyramid, 40 HU noise threshold.

Patients 1, 2, 4, and 5 produced high-quality dual-phase composites with clear bilateral renal enhancement using the coarse-register/fine-apply strategy. Patient 3 was processed in venous-only mode due to the non-uniform arterial slice spacing (23.6 mm maximum gap) producing RIFE interpolation artifacts. Full-resolution 1.0 mm registration (16.5 minutes) was required for Patient 3 because the coarse strategy depended on the same non-uniform data. The venous phase covered 48.2% of the noncontrast Z-axis extent (340 of 705 slices); the Z-axis validity mask correctly zeroed enhancement outside this region. Mean venous enhancement in Patient 3 was 347 HU, confirming adequate registration quality despite the data limitations.

## 6. Discussion

Beyond PRISM itself, our experiments highlight general lessons for high-resolution deformable registration pipelines:

- **Optimizer budget vs. dimensionality**. Finer B-spline grids introduce many more parameters; without a proportional increase in optimizer evaluations or an adaptive stopping criterion, the optimizer may terminate before exploiting their theoretical accuracy advantage.
- **Pyramid scale vs. grid spacing**. Multi-resolution pyramid design must be matched to the spatial scale of the deformation model; when B-spline control points are spaced on the order of tens of millimeters, additional fine-resolution pyramid levels add substantial cost without measurable gains in alignment.
- **Coarse scales vs. voxel resolution**. In abdominal CT, the effective deformation scale is much coarser than the voxel grid, so registration quality can remain stable across a wide range of voxel spacings, enabling coarse-register/fine-apply strategies.

The most consequential finding of this parameter optimization study is the independence of B-spline registration quality from both metric sampling fraction (Experiment B) and voxel resolution (Experiment D). These two results together validate the coarse-register/fine-apply strategy, reducing dual-phase study registration from approximately 20 minutes to 8 minutes while preserving sub-millimeter spatial detail. With the complete optimized configuration, PRISM delivers 0.5 mm registered DSA volumes in approximately 14 minutes on a single GPU, a clinically feasible throughput for retrospective cohort analysis or batch processing.

The failure of the finest grid spacing (20 mm) to outperform 30 mm (Experiment A) highlights a fundamental principle for limited-budget optimizers in high-dimensional spaces: the optimizer evaluation budget must scale with the parameter space dimensionality. With 38,603 parameters and only 100 function evaluations, the L-BFGS-B gradient tolerance criterion was satisfied before meaningful optimization could occur for the largest patient (Patient 3, 60,306 parameters). Future work will investigate scaling the function evaluation limit with the square root of parameter count, or by using adaptive line-search and early-stopping heuristics tailored to MI landscapes. Increasing the evaluation budget to 500–1000 iterations would likely allow finer grids to realize their theoretical quality advantage.

The failure of the [4,2] two-level pyramid to improve over [4] (Experiment C) is consistent with the spatial scale argument: a 40 mm B-spline grid captures body-scale deformations that are fully represented at 4× downsampling. The additional 2× refinement level adds no information because there are no genuine tissue deformations at spatial frequencies above the 40 mm control point spacing that rigid and affine steps have not already corrected. This principle likely generalizes: the choice of pyramid configuration should be guided by the B-spline grid spacing, with finer grids (10–20 mm, capturing tumor-scale deformations) potentially benefiting from additional refinement levels.

Patient 3 consistently underperformed across all experiments, illustrating the critical role of data quality gates in multi-phase CT registration pipelines. Non-uniform slice spacing introduced RIFE interpolation artifacts (not previously known), while an incompatible field-of-view position required manual series replacement. Automated preprocessing checks such as slice spacing uniformity assessment, field-of-view compatibility verification, and contrast timing validation are therefore essential components of a robust production pipeline. Excluding Patient 3 would increase mean MI improvements by approximately 15–20% across all experiments.

Some limitations warrant acknowledgment. First, the sample size of five patients limits statistical power; results should be interpreted as engineering-focused parameter optimization guidance rather than clinical performance estimates. Second, registration quality was assessed using Mattes Mutual Information only; landmark-based or segmentation-overlap metrics may reveal quality differences not captured by MI. Third, the L-BFGS-B optimizer was limited to 100 function evaluations across all experiments, which systematically disadvantaged fine grid spacings. Fourth, the coarse-register/fine-apply strategy assumes the B-spline transform computed at 3.0 mm accurately represents tissue deformation at finer scales; highly localized deformations in small tumors with mass effect may benefit from direct fine-resolution registration. Future work could apply fine-grid, high-budget optimization on a subset of patients to validate the coarse strategy.

## 7. Conclusion

We present PRISM, an open-source automated pipeline for multi-phase CT digital subtraction angiography, alongside a systematic parameter optimization study establishing evidence-based configuration recommendations for B-spline deformable registration in high-resolution abdominal CT. The five-experiment benchmark (200 registration runs; ~25 GPU-hours) identified a consistent set of optimal parameters: 40 mm B-spline grid spacing, 5% metric sampling, single-level [4] pyramid, 0.5 mm interpolation target with coarse-register/fine-apply, and 40 HU subtraction noise threshold. The finding that registration quality is independent of voxel resolution across 0.5–3.0 mm motivates the coarse-register/fine-apply strategy, reducing dual-phase registration time by approximately 60% while preserving sub-millimeter spatial detail.

PRISM is publicly available at github.com/rrodrig30/prism. Prospective multi-institutional validation with radiologist readers remains the critical next step toward establishing PRISM-generated DSA as a diagnostic adjunct for renal mass characterization. Beyond renal imaging, the parameter configurations we identify for grid spacing, sampling, pyramid design, and voxel resolution provide practical guidance for designing efficient deformable registration pipelines in other high-resolution CT applications.

## Data Availability

The PRISM source code is openly available at https://github.com/rrodrig30/prism. The clinical multi-phase CT datasets analyzed in this study are not publicly available because they contain protected health information and their release is restricted by institutional policy; de-identified derived quantitative results (registration timings, MI improvement values, and subtraction metrics for all 200 registration runs) are available from the corresponding author upon reasonable request.

## Acknowledgments

The authors thank the developers of SimpleITK, TotalSegmentator, RIFE, nnU-Net, and the KiTS challenge organizers for making their tools publicly available. Computational resources were provided by a NVIDIA H100 NVL GPU system. This work was supported by grants from the Abraham and Linda Littenberg Foundation and the Deborah and Wayne Staysniak Foundation.

## Supplemental Data

### A: Software Architecture

PRISM is implemented as a modular Python web application comprising approximately 45 source files organized into six packages: core, imaging, reconstruction, web, db, and utils. The system enforces strict dependency layering where lower-level packages (utils, db) never import from higher-level packages, ensuring clean separation of concerns and testability.

The processing pipeline is managed by a PipelineController that maintains state through a PipelineStage enumeration (integer values 0–6). Each stage verifies that its predecessor has completed successfully before permitting execution and supports rollback to any previous stage. Progress is streamed to the browser via HTMX polling with self-replacing HTML fragments.

#### Technology stack

The backend is built on FastAPI (Ramírez, 2019) with Uvicorn for asynchronous request handling. Long-running pipeline operations execute in background threads via asyncio.to_thread(), with progress tracked through module-level job dictionaries. The frontend uses server-rendered Jinja2 templates enhanced with HTMX for partial page updates and Alpine.js for client-side interactivity. Two-dimensional slice viewing uses Cornerstone.js; three-dimensional visualization uses VTK.js on the client with PyVista for server-side mesh generation.

#### Thread management

ITK defaults to spawning threads equal to the number of available CPU cores. On the 76-core test system, this caused aggregate memory consumption during B-spline optimization to reach 878 GB before out-of-memory termination. Explicitly limiting ITK threads to 16 via sitk.ProcessObject.SetGlobalDefaultNumberOfThreads(16) reduced peak memory to 14.8 GB (59× reduction) with minimal impact on wall-clock time.

#### Deployment

All runtime parameters are loaded from environment variables with a PRISM_ prefix. Docker deployment is supported through a compose configuration with NVIDIA runtime for GPU passthrough.

### B: RIFE Padding Defect and Correction

An initial version of the RIFE interpolation engine padded input slices to multiples of 32 pixels. This produced correct results for most input dimensions but caused tensor size mismatches in IFNet-HDv3 at certain resolutions (notably 0.5 mm target spacing with 3+ mm source spacing). The root cause is that IFNet-HDv3 has five scale levels that progressively halve feature maps; when an input dimension is an odd multiple of 32 (e.g., 928 = 29 × 32), intermediate feature maps at coarser levels have fractional sizes that do not match across the encoder–decoder pathway. The fix is to pad to multiples of 128 (= 2^7^), guaranteeing clean division through all five scale levels. This correction enabled the 0.5 mm results reported in Experiment D and has been incorporated into the production PRISM pipeline.

### C: Data Quality Gates for Multi-Phase CT

Experience with the five-patient cohort—particularly Patient 3—identified three critical preprocessing quality checks for multi-phase CT registration pipelines:

#### Slice spacing uniformity

Maximum inter-slice gap should not exceed 2× the nominal slice thickness. Non-uniform gaps (e.g., 23.6 mm in Patient 3’s arterial series) produce RIFE interpolation artifacts that generate spurious enhancement signal across the entire subtraction volume.

#### Field-of-view compatibility

The physical X/Y/Z origin coordinates of each contrast phase must overlap with the noncontrast reference. An offset exceeding approximately 50 mm typically indicates a non-axial reconstruction or an incorrect series selection. Resampling a completely non-overlapping volume to the reference grid produces a background-filled (−1000 HU) volume.

#### Contrast phase ordering

When multiple candidate series exist for a given phase, field-of-view compatibility should be verified before accepting the series identified by the phase classification algorithm. The phase classifier optimizes for temporal and histogram features; it does not verify spatial compatibility.

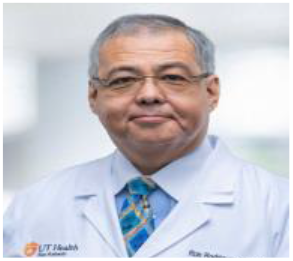

**Ronald Rodriguez, MD, PhD** is a urologic oncologist and professor in the Department of Urology and Medical Education at UT Health San Antonio, San Antonio, Texas, USA. His research focuses on renal cell carcinoma, image-guided diagnosis and therapy, and the integration of advanced imaging techniques into clinical urologic oncology workflows.

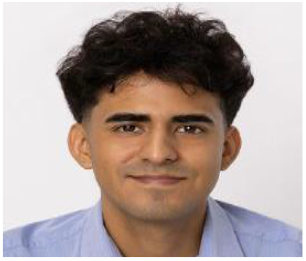

**Ramiro Ramirez, MD** is a radiologist and educator in the Department of Urology and Medical Education at UT Health San Antonio, San Antonio, Texas, USA. His interests include abdominal and genitourinary imaging, CT angiography, and the development of practical imaging pipelines that can be translated into routine clinical practice.

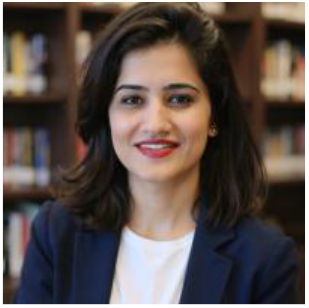

**Heena Rathore, PhD** is an assistant professor in the Department of Computer Science at Texas State University, San Marcos, Texas, USA. Her research spans medical image analysis, machine learning systems, and high-performance computing, with a focus on deploying robust, interpretable AI pipelines for clinical imaging applications.

## Notes

### Competing Interest Statement

The authors have declared no competing interest.

### Author Declarations

The Institutional Review Board of The University of Texas Health Science Center at San Antonio gave ethical approval for this work.

## References

Filho, A. M., M. Laversanne, J. Ferlay, M. Colombet, M. Pineros, A. Znaor, D. M. Parkin, I. Soerjomataram, and F. Bray. 2025. ‘The GLOBOCAN 2022 cancer estimates: Data sources, methods, and a snapshot of the cancer burden worldwide’, Int J Cancer, 156: 1336–46.

Gambini, L., C. Gabbett, L. Doolan, L. Jones, J. N. Coleman, P. Gilligan, and S. Sanvito. 2024. ‘Video frame interpolation neural network for 3D tomography across different length scales’, Nat Commun, 15: 7962.

Gan, X., S. Zhu, Y. Zhang, Z. Wang, X. Ye, K. Hu, and X. Gao. 2026. ‘Dual-perspective decoupling network for kidney tumor segmentation on CT images’, Neural Netw, 193: 108042.

Heller, N., F. Isensee, K. H. Maier-Hein, X. Hou, C. Xie, F. Li, Y. Nan, G. Mu, Z. Lin, M. Han, G. Yao, Y. Gao, Y. Zhang, Y. Wang, F. Hou, J. Yang, G. Xiong, J. Tian, C. Zhong, J. Ma, J. Rickman, J. Dean, B. Stai, R. Tejpaul, M. Oestreich, P. Blake, H. Kaluzniak, S. Raza, J. Rosenberg, K. Moore, E. Walczak, Z. Rengel, Z. Edgerton, R. Vasdev, M. Peterson, S. McSweeney, S. Peterson, A. Kalapara, N. Sathianathen, N. Papanikolopoulos, and C. Weight. 2021. ‘The state of the art in kidney and kidney tumor segmentation in contrast-enhanced CT imaging: Results of the KiTS19 challenge’, Med Image Anal, 67: 101821.

Heller, N., Fabian Isensee, D. A. Trofimova, Resha Tejpaul, Zhongchen Zhao, Huai Chen, Lisheng Wang, Alexander M. Golts, Daniel Khapun, Daniel Shats, Yoel Shoshan, Flora Gilboa-Solomon, Yasmeen M. George, Xi Yang, Jianpeng Zhang, Jing Zhang, Yong Xia, Mengran Wu, Zhiyang Liu, Edward Walczak, Sean McSweeney, Ranveer M.S. Vasdev, Christopher M. Hornung, Rafat H. Solaiman, Jamee Schoephoerster, Bailey R. Abernathy, Davi Chen Wu, Safa Abdulkadir, Benjamin C. Byun, Justice Spriggs, Griffin Struyk, Alexandra Austin, B. Simpson, Michael Hagstrom, Sierra Virnig, John M French, N. Venkatesh, Sarah Cheuk Hei Chan, Keenan Moore, Anna Jacobsen, Susan Austin, M. Austin, Subodh K. Regmi, Nikolaos Papanikolopoulos, and Christopher J. Weight. 2023. ‘The KiTS21 Challenge: Automatic segmentation of kidneys, renal tumors, and renal cysts in corticomedullary-phase CT’, arXiv, abs/2307.01984.

Huang, Z., T. Zhang, W. Heng, B. Shi, and S. Zhou. 2022. “Real-time intermediate flow estimation for video frame interpolation.” In Proceedings of the European Conference on Computer Vision (ECCV), 624–42. Springer.

Isensee, F., P. F. Jaeger, S. A. A. Kohl, J. Petersen, and K. H. Maier-Hein. 2021. ‘nnU-Net: a self-configuring method for deep learning-based biomedical image segmentation’, Nat Methods, 18: 203–11.

Israel, G. M., and M. A. Bosniak. 2005. ‘How I do it: evaluating renal masses’, Radiology, 236: 441–50.

Kato, Y., M. Hayakawa, H. Sano, M. V. Sunil, S. Imizu, M. Yoneda, S. Watanabe, M. Abe, and T. Kanno. 2004. ‘Prediction of impending rupture in aneurysms using 4D-CTA: histopathological verification of a real-time minimally invasive tool in unruptured aneurysms’, Minim Invasive Neurosurg, 47: 131–5.

Kim, J. K., T. K. Kim, H. J. Ahn, C. S. Kim, K. R. Kim, and K. S. Cho. 2002. ‘Differentiation of subtypes of renal cell carcinoma on helical CT scans’, AJR Am J Roentgenol, 178: 1499–506.

Koetzier, L. R., D. Mastrodicasa, T. P. Szczykutowicz, N. R. van der Werf, A. S. Wang, V. Sandfort, A. J. van der Molen, D. Fleischmann, and M. J. Willemink. 2023. ‘Deep Learning Image Reconstruction for CT: Technical Principles and Clinical Prospects’, Radiology, 306: e221257.

Kotre, C. J., and N. W. Marshall. 2001. ‘A review of image quality and dose issues in digital fluorography and digital subtraction angiography’, Radiat Prot Dosimetry, 94: 73–6.

Lowekamp, B. C., D. T. Chen, L. Ibanez, and D. Blezek. 2013. ‘The Design of SimpleITK’, Front Neuroinform, 7: 45.

Ma, G., Y. Yu, H. Duan, Y. Dou, Y. Jia, X. Zhang, C. Yang, X. Chen, D. Han, C. Guo, and T. He. 2018. ‘Subtraction CT angiography in head and neck with low radiation and contrast dose dual-energy spectral CT using rapid kV-switching technique’, Br J Radiol, 91: 20170631.

Ramírez, S. 2019. “FastAPI: Modern, fast (high-performance) web framework for building APIs with Python.” In.

Reis, E. P., L. Blankemeier, J. M. Zambrano Chaves, M. E. K. Jensen, S. Yao, C. A. M. Truyts, M. H. Willis, S. Adams, E. Amaro, Jr., R. D. Boutin, and A. S. Chaudhari. 2024. ‘Automated abdominal CT contrast phase detection using an interpretable and open-source artificial intelligence algorithm’, Eur Radiol, 34: 6680–87.

Rueckert, D., L. I. Sonoda, C. Hayes, D. L. Hill, M. O. Leach, and D. J. Hawkes. 1999. ‘Nonrigid registration using free-form deformations: application to breast MR images’, IEEE Trans Med Imaging, 18: 712–21.

Siegel, R. L., T. B. Kratzer, N. S. Wagle, H. Sung, and A. Jemal. 2026. ‘Cancer statistics, 2026’, CA Cancer J Clin, 76: e70043.

Te Kiefte, B. J. C., F. Gholamiankhah, J. F. Juffermans, P. Van Den Boogaard, Ajha Scholte, H. J. Lamb, and J. J. M. Westenberg. 2025. ‘Multimodality comparison of aorta morphology in patients with aortopathy: 4D flow CMR, CTA, mDIXON’, BMC Med Imaging, 25: 201.

Thomas, C., A. Korn, B. Krauss, D. Ketelsen, I. Tsiflikas, A. Reimann, H. Brodoefel, C. D. Claussen, A. F. Kopp, U. Ernemann, and M. Heuschmid. 2010. ‘Automatic bone and plaque removal using dual energy CT for head and neck angiography: feasibility and initial performance evaluation’, Eur J Radiol, 76: 61–7.

Wasserthal, J., H. C. Breit, M. T. Meyer, M. Pradella, D. Hinck, A. W. Sauter, T. Heye, D. T. Boll, J. Cyriac, S. Yang, M. Bach, and M. Segeroth. 2023. ‘TotalSegmentator: Robust Segmentation of 104 Anatomic Structures in CT Images’, Radiol Artif Intell, 5: e230024.

Wu, Z., J. Wei, J. Wang, and R. Li. 2022. ‘Slice imputation: Multiple intermediate slices interpolation for anisotropic 3D medical image segmentation’, Comput Biol Med, 147: 105667.

